# The Effects of Indoor Residual Spraying on Prevalence of Malaria among Under-five Children in Zambia; A Retrospective Cohort Study

**DOI:** 10.1101/2024.05.27.24307974

**Authors:** Gwenie Kapiya, Cephas Sialubanje, Mukumbuta Nawa

## Abstract

**Introduction:** Malaria remains a global challenge despite the efforts to eliminate it by 2030 by the WHO and its partner countries. About 93.6% of the malaria cases and 95.4% of the deaths occurred in Africa. Zambia is one of the top 20 highly endemic countries with about a third (29.3%) of all children aged 6 to 59 months having malaria in 2021 and it accounts for about 1.4% of the global malaria incidence and mortality. Among the interventions used to eliminate malaria is Indoor Residual Spraying (IRS). Existing literature has shown the effects of IRS on malaria prevalence and incidence by comparing IRS versus no IRS intervention. This study assessed the effects of IRS on malaria prevalence over time from when it was done in three monthly cohorts over a period of one year.

**Methods:** This study was a retrospective cohort study. Data was collected retrospectively covering a period of 12 months when the IRS was done in the households where the Malaria Indicator Survey of 2021 was carried out. The study then compared malaria prevalence in closed cohorts of three months. Data was analysed in Stata version 14, descriptive statistics were summarized as counts and percentages, cross-tabulations between the dependent variable and independent variables were done and measures of association were assessed using univariate and multivariable logistic regression. The level of significance was set at 0.05.

**Results:** The study included 1,786 children aged six to 59 months and more of these were female 52.5% (938/1786). Among the children, 14.7% (263/1786) were in the 0 to 3 months cohort, 59.3% (1059/1786) were in the 4 to 6 months cohort, 16.2% (289/1786) in the 7 to 9 months cohort and 9.8% (175/1786) in the 10 to 12 months cohort. The malaria prevalence was 33.4%. There were no significant statistical differences in the malaria prevalence among children in the four cohorts.

**Conclusion:** This study found that over a period of one year, the protective effect of Indoor Residual Spraying was not statistically different among under-five children whether it was done within zero to three months, four to six months, seven to nine months or ten to twelve months. This implies that the protective effects of IRS on malaria prevalence extended equally over a period of 12 months.

## INTRODUCTION

Malaria is a parasitic disease that is spread through bites of the female anopheles mosquito (1). It affects mostly under-five children and pregnant women (2). The current global estimates of malaria indicate that there were 249 million (95%CI 225 – 278 million) cases and 608,000 (95%CI 566,000 – 738,000) deaths attributable to malaria in 2022 (3). About 93.6% of the malaria cases and 95.4% of the deaths occurred in Africa (3). Zambia is one of the top 20 highly endemic countries in the world with about a third (29.3%) of all children aged below five years having malaria in 2021 and accounting for 1.4% of the global malaria incidence and mortality (4, 5).

Zambia through its Ministry of Health’s National Malaria Elimination Program aims to eliminate malaria by the year 2030 and interrupt the reestablishment of local transmission of malaria thereafter (6). The interventions to eliminate malaria include multifaceted strategies such as vector control through Indoor Residual Spraying (IRS), distribution of Insecticide Treated Nets (ITNs), larviciding, prompt facility and community diagnosis through the use of Rapid Diagnostic Tests (RDTs) and treatment using Artemisinin Combination Therapies (ACTs), routine antimalarial prophylaxis in pregnant women, surveillance, mass drug administration in selected areas, malaria vaccine, active and passive case detection (6).

The use of Indoor Residual Spraying has been documented to be an effective strategy in fighting malaria (7). IRS chemicals remain active for six to twelve months and can prevent mosquito vectors from biting people because the main malaria-transmitting mosquito vectors in Africa prefer indoors for resting and biting (7). However, the IRS has not been universally deployed in Zambia because of some logistical challenges such as poor quality of housing structures which are not suitable for the IRS, high cost of chemicals, deployment in procurement procedures of chemicals, lack of transport, emergency of resistance to the chemicals and the changing of mosquito behaviour to feeding at dusk and dawn (8). Despite these challenges, the IRS remains one of the backbones of malaria prevention in Zambia together with the use of ITNs (6). Recent entomological studies on the effects of IRS in high transmission areas in Zambia have shown a 40 – 50% decrease in the indoor abundance of primary vectors such as *Anopheles gambiae* and *funestus* (9). A recent systematic review and meta-analysis on the effects of IRS that included over 1.1 million people in IRS-sprayed households found that the odds of malaria infection were lowered at 0.35 (95%CI 0.27 – 0.44) compared to non-sprayed households (10). A study done in Zambia found that IRS significantly reduced the incidence of malaria IRR 0.91 (95%CI 0.84 – 0.98) in sprayed areas compared to areas that were not sprayed with pirimiphos methyl (11). The effects of the IRS despite being designed to last for long periods, wanes over time, which necessitates repeats of applications to ensure continued effectiveness. A recent study in Uganda found that the periods of effectiveness were dependent on the type of chemicals used, for bendiocarb, a carbamate type of chemical, the period was 17 weeks (95%CI 14 – 21.) whilst pirimiphos methyl an organophosphate, the time of effectiveness was longer at 40 weeks (95%CI 37– 42) (12). Given this time dependency of the optimal effectiveness of IRS on malaria infection, this study assessed the malaria prevalence in different cohorts of under-five children in Zambia. Studies that assess the effectiveness of the IRS over time are usually in controlled environments and closely monitored. Under field conditions in a high malaria endemic country such as Zambia, we do not know the effects of IRS over time within one year. We therefore used data collected from a nationwide cross-sectional survey the Malaria Indicator Survey (MIS) and divided households based on the months back when they were sprayed into three months cohorts of zero to three, four to six, seven to nine and ten to twelve months cohorts. Children in those households were tested for malaria using rapid diagnostic tests and we compared malaria prevalence ratios among the cohorts after adjusting for other confounders.

### Conceptual Framework

Multiple factors influence the effects of IRS on malaria prevalence and incidence which include the type of chemicals that are used. Some chemicals have a longer time of effectiveness and others have short timespans. The coverage of the IRS is also important, studies have shown that at least 80% coverage of households is more effective than below 80% (13). In addition, the species of malaria mosquito vectors and their behaviours affect the effects of IRS on malaria prevention. IRS is most effective in Africa where predominant species such as *Anopheles gambiae* and *funestu*s are highly anthropophilic and endophillic (14). When primary vectors are exophagic and zoophilic, indoor-based interventions such as IRS and ITNs may not be effective (15). Further, environmental factors such as drought, flooding, temperature and rainfall affect the population and species of vector mosquitoes in an area. Other interventions like ITN deployment also affect the effectiveness of the IRS. Socioeconomic factors such as household income, type of house and nutrition are other factors that should be explored when assessing the effect of interventions such as IRS.

#### IRS Coverage and Types of Chemicals

Indoor Residual Spraying has been deployed in Zambia for more than twenty years alongside other interventions like ITNs (16). As of 2021, IRS was carried out in 115 districts out of the 116 districts in Zambia and a total of 2, 720,479 structures were sprayed (4). The percentage of households sprayed was 39% and the average duration from the time of spraying was seven months (4). The types of chemicals used for spraying in Zambia are those to which the primary vectors are not resistant (17). Zambia currently uses World Health Organization Pesticide Evaluation Scheme (WHOPES) approved insecticides such as Pirimiphos Methyl and Fludora Fusion a mixture of deltamethrin and clothanidine (17, 18). Both Pirimiphos Methyl and Fludora Fusion have long periods of effectiveness ranging from 8 – 12 months, although mortality rates have been noted to reduce from greater than 80% up to eight months and decline by 50% after eight months (18). There are five groups of chemicals that have been used for IRS including pyrethroids, carbamates, organophosphates, organochlorines and neonicotinoids. In Zambia, countrywide entomological studies have demonstrated high resistance to some pyrethroids and carbamate groups of chemicals and are therefore not used for IRS activities except in combinations with synergists that negate the resistance (19).

### Socioeconomic Factors

Socioeconomic factors such as residence in rural areas, occupation in farming and occupations that expose people to mosquito bites influence malaria transmission (20). Economic activities such as keeping cattle, goats and chicken have also been associated with increased effects of mosquito bites and malaria transmission risk (20). Even when your house has been sprayed with IRS chemicals, if your occupation exposes you to mosquito bites, the protective effects of IRS when you are sleeping at home will not be of effect. Other economic factors include the type of housing that people can afford because of income levels and social status, modern houses have been shown to be insect-proof and protective against malaria transmission compared to traditional houses in Africa which are made of grass and mud and allow mosquitoes to enter the house freely (21). Further, the application of IRS chemicals is easier and lasts longer in modern houses that are coated with paints as opposed to natural materials such as grass or mud. In fact, in Zambia, before applying IRS, the programs assess the suitability of the housing structures in line with recommendations for IRS applications (6). Some poor-quality structures of grass and mud are not suitable for IRS application (6). Other socioeconomic variables include wealth status, children from lower income brackets are more likely to have malaria due to other predisposing factors such as poor nutrition, anaemia, and lack of access to interventions [6].

### Species of Malaria Mosquito Vectors

Out of the over 450 species of Anopheline mosquitoes, about 90 species are known to transmit malaria to humans, among these, an even limited number of species have the capacity and competence to sustain local transmission within specific areas (22). Moreover, in Zambia, the main primary mosquito vectors are *An. gambiae sensu stricto* Giles, *Anopheles arabiensis* Paton, and *Anopheles funestus sensu stricto* Giles (19). These vectors are known to be anthropophagic, endophagic and endophillic, this makes indoor interventions such as IRS and ITN effective in fighting malaria (23). *An. arabiensis* has been noted to exhibit plasticity in host preferences, whilst some secondary vectors such as *An. rufipes, An. squamosus* and *An. coustani* were largely zoophilic and exophagic (17, 24). In most areas, many species of mosquito coexist in sympatry and allopatric speciation (25). Therefore, while indoor-based interventions such as IRS and ITNs are good in addressing transmission from the main vectors which are endophagic and endophillic, some species such as *An. arabiensis* and some secondary vectors have exhibited crepuscular behaviour, exophagic and zoophilic, therefore they can bite in the evenings when people are outside or at dusk or when they go out for some field activities [24]. To address these, there is a need for additional interventions such as mosquito repellents and vaccines (17).

### Environmental Factors

A vector-borne disease such as malaria is largely influenced by environmental factors such as rainfall, temperature, presence of large water bodies and vegetation cover. The temperatures in sub-Saharan Africa and Zambia in particular are conducive to mosquito vector breeding (26). The development of the aquatic stages of the vectors is dependent on the ambient temperatures particularly the minimum temperatures (26). Temperatures below 16 degrees Celsius and those above 32 degrees Celsius are not very suitable whilst between 16 – 32 are suitable with optimum at around 25 degrees (26). Warmer temperatures within the suitability range enhance the development whilst lower temperatures delay the development (27). Very hot temperatures also affect the foraging behaviour and longevity of the adult mosquitoes as they kill them and mosquitoes have been known to become less active and hide until conditions are suitable, this state of inactivity during dry hot spells is known as aestivation (28). When rains come, and the temperatures become suitable, the adults emerge and begin foraging and reproducing to replenish the populations of mosquito vectors (28). During winters with extremely low temperatures, adult mosquitoes are also known to hibernate in cellars and other above-ground structures where they hide, lower their basal metabolic rates and can survive for longer periods until conditions improve (29).

Rainfall affects the suitability of malaria mosquito vectors, absence of rainfall such as prolonged droughts would reduce mosquito populations and would lead some species to estivate until conditions improve (28). On the other hand, abundant rainfall and humidity are conducive for mosquito breeding leading to high mosquito populations and transmission of malaria. In Zambia, the northern parts of the country near the equator receive higher rainfall, have larger water bodies and have higher transmission of malaria compared to the drier parts in Southern and Western provinces which are nearer to the Kalahari and Namib deserts and have low transmission of malaria (20).

## METHODOLOGY

### Study Design

This study is going to be a retrospective cohort study. Data will be collected retrospectively covering a period of 12 months on when IRS was done in the households where the Malaria Indicator Survey of 2021 was carried out. The study will then compare malaria prevalence in closed cohorts of three months.

### Study Settings

The study was carried out in all ten provinces of Zambia and powered to estimate malaria nationally, at the provincial level and in the urban and rural strata. Zambia is a southern African country with a tropical climate and has malaria transmission throughout the year with peak malaria season during the rainy and hot season from November to April (30). The country then experiences a dry cool season from May to August and then from September to October, the weather is hot and dry (31). Zambia experience more malaria in the northern parts of the country which receive more rainfall than the southern parts which receive less rainfall (31).

### Study Population

This study was carried out in children aged six to 59 months.

### 4.4 Sample Size Determination

This study will include all under-five children who were included in the database for the period of the data collection. The minimum dataset was calculated at 95% confidence level, 80% power and an estimated malaria prevalence of 29.3%. In the different cohorts, we powered the study to detect at least a 10% difference.

N = (p_0_q_0_ + p_1_q_1_) (z_α/2_ + z_β_)^2^.df / (p_1_ – p_0_)^2^

Where n = number of respondents

.p0 = proportion of respondents with no exposure = 29.3% malaria (4).
.p1 = proportion of respondents with exposure = 10% difference
.α = level of significance = 0.05
.β = power = 80%
.df = design effect = 1.5

This study therefore needs a minimum of 1,125 respondents, however, because it will use secondary data, all under-five children who were included in the primary study will be included.

### 4.5 Data Collection and Analysis

The data was extracted from the database at the National Malaria Elimination Programme of the Ministry of Health using a Microsoft Comma Delimited file. This was imported into Stata version 14 where it was cleaned and validated. It was then analysed using descriptive statistics such as counts, frequencies, percentages, mean, median, confidence intervals and interquartile range. Comparisons were made using cross-tabulations and hypothesis tests conducted using chi-square, or Fisher’s exact test. Further, comparisons using prevalence ratios were done using univariate and multivariable Poisson regression. The level of significance was set at 0.05.

### 4.6 Ethical Considerations

Permission was obtained from the Ministry of Health to access the dataset at the National Malaria Elimination Program (MNEP). Ethical review was obtained from the Lusaka Apex Medical University Ethical Review Committee. Permission was obtained from the National Health Research Authority to carry out this study. Further, the confidentiality of the data was assured through the anonymity of the dataset. Security of the data was assured through password protection and back on external media that was password protected. Informed consent was already obtained from the guardians of the children when the primary data was being collected.

## RESULTS

### Socio-demographic and Prevalence of Malaria and Anaemia

A total of 1,786 children below the age of five years were included in this study, this was more than the 1,125 that was required based on the sample size calculation. There were more females 938/1786 (52.5%) than males 848/1786 (47.5%) which was a reflection of the general population in Zambia where there are more females than males. Fewer children were below one year compared to 227/1786 (12.7%) while those one to four years were close to 20% each year. The majority of the heads of the households where the children resided had no or primary education 59.5% and very few had tertiary education 2.9%. The majority were from rural areas 91.1% and all the respondents had their house sprayed as this was a cohort study of indoor residual spraying (IRS). The malaria prevalence in this one-year cohort was 33.4% whilst anaemia prevalence was 46.6%. Table 1 summarises the basic characteristics and prevalence of malaria, anaemia and fever among the respondents.

**Figure 1:**
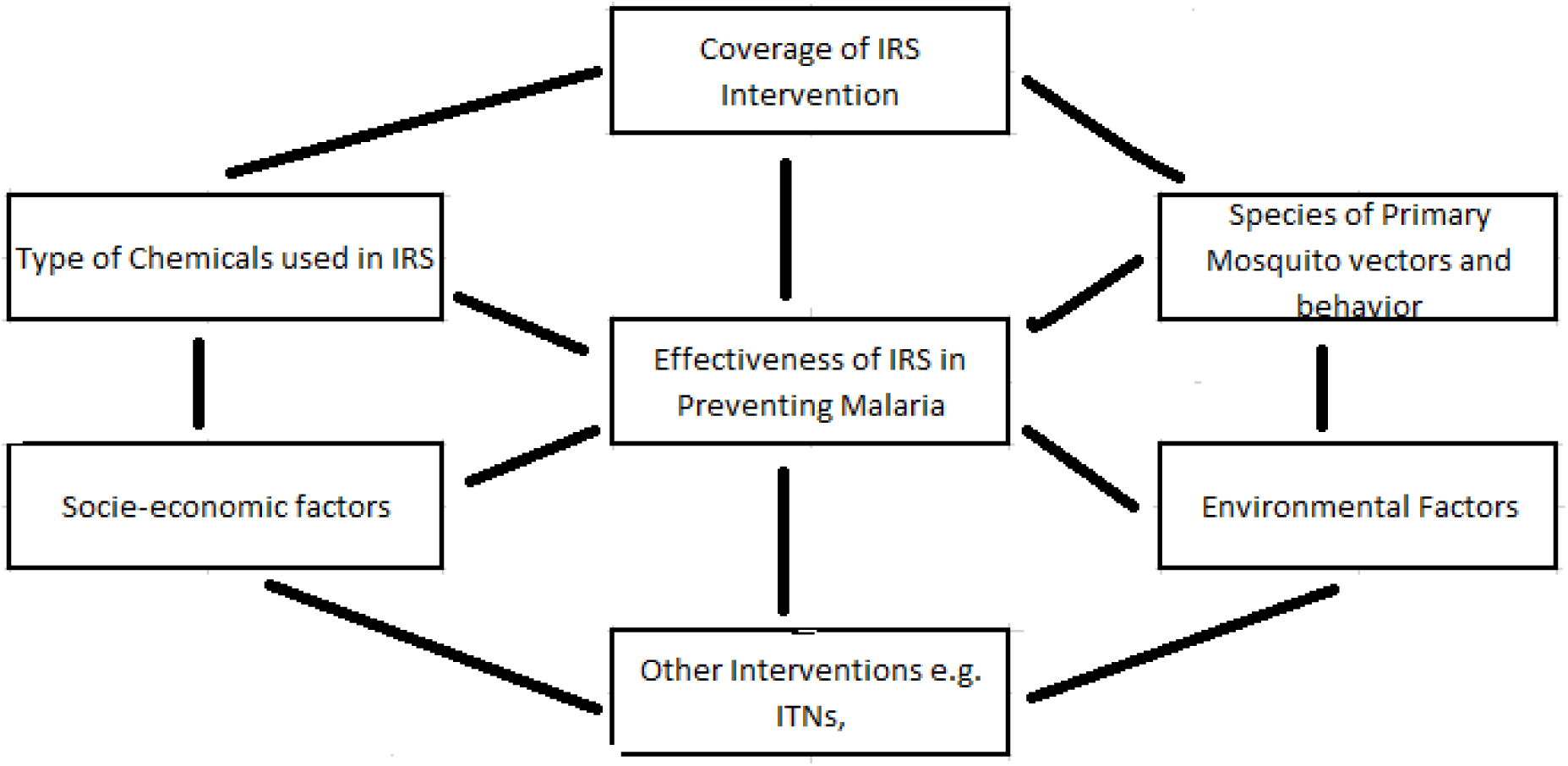
Factors affecting the Effects of IRS on Malaria; Source: Authors.

**Table 1:** Basic Characteristics, Malaria, Anaemia and Fever Prevalence.

| Variable | Categories | No. | Percentage | P value |
| --- | --- | --- | --- | --- |
| Sex | Female | 938 | 52.5 | **0.033\*** |
|  | Male | 848 | 47.5 |  |
| Age (Years) | Below 1 Year | 227 | 12.7 | **< 0.001\*\*** |
|  | One year | 341 | 19.1 |  |
|  | Two year | 390 | 21.8 |  |
|  | Three Years | 370 | 20.8 |  |
|  | Four Years | 458 | 25.6 |  |
| Education Household Head | No or Primary | 869 | 59.5 | **< 0.001\*\*** |
|  | Secondary | 550 | 37.7 |  |
|  | Tertiary | 42 | 2.9 |  |
| Residence Type | Rural | 1627 | 91.1 | **< 0.001\*\*** |
|  | Urban | 159 | 8.9 |  |
| Province | Central | 90 | 5 | **< 0.001\*\*** |
|  | Copperbelt | 204 | 11.4 |  |
|  | Eastern | 525 | 29.4 |  |
|  | Luapula | 290 | 16.2 |  |
|  | Lusaka | 65 | 3.6 |  |
|  | Muchinga | 157 | 7.1 |  |
|  | Northern | 152 | 8.5 |  |
|  | North-Western | 96 | 5.4 |  |
|  | Southern | 87 | 4.9 |  |
|  | Western | 150 | 8.4 |  |
| Wealth Quintiles | 1 (Lowest) | 427 | 23.9 | **< 0.001\*\*** |
|  | 2 (Upper Low) | 349 | 19.5 |  |
|  | 3 (Middle) | 382 | 21.4 |  |
|  | 4 (Lower High) | 421 | 23.6 |  |
|  | 5 (Highest) | 207 | 11.6 |  |
| House Indoor Residual Spray | Sprayed | 1786 | 100 | **< 0.001\*** |
|  | Not Sprayed | 0 | 0 |  |
| Months Ago Sprayed (IRS) | 0 - 3 Months | 263 | 14.7 | **< 0.001** |
|  | 4 - 6 Months | 1059 | 59.3 |  |
|  | 7 - 9 Months | 289 | 16.2 |  |
|  | 10 - 12 Months | 175 | 9.8 |  |
| ITN Use | Slept ITN | 753 | 82.7 | < 0.001* |
|  | Not Slept ITN | 158 | 17.3 |  |
| Malaria (RDT) | Positive | 1189 | 66.6 | < 0.001* |
|  | Negative | 597 | 33.4 |  |
| Fever in Last two weeks | Fever | 420 | 23.5 | < 0.001* |
|  | No Fever | 1366 | 76.5 |  |
| Haemoglobin | Anaemic | 923 | 53.4 | < 0.005* |
|  | Not Anaemic | 806 | 46.6 |  |

\* Test of Proportions
\*\* Multiple Proportions Test (Marascuilo Procedure)

### Malaria by Different Characteristics

In the preliminary analysis, Table 2 shows malaria prevalence stratified by different characteristics, it was not different between males and females (P-value = 0.805), however, it was different in all other variables such as age, rural or urban residence, province, wealth quintiles, ITN use, haemoglobin and fever. Specifically, in the stratified IRS cohorts, malaria was highest in the 3-6 months cohort at 38.4% and second in the 7 – 9 months cohort at 30.8%. These findings may suggest that malaria is lower in the 10 – 12 months cohort but the findings are still confounded and were further adjusted in the Poisson multivariable regression model.

**Table 2:** Malaria Prevalence Stratified by Different Variables.

| Variable | Categories | Rapid Diagnostic Test Result |  | P-Value |
| --- | --- | --- | --- | --- |
|  |  | No Malaria n(%) | Malaria n(%) |  |
| Sex | Female | 622 (66.3) | 316 (33.7) | 0.805* |
|  | Male | 567 (66.9) | 281 (33.1) |  |
| Age (Years) | Below 1 Year | 176 (77.5) | 51 (22.5) | < 0.001* |
|  | One year | 259 (76.0) | 82 (24.0) |  |
|  | Two year | 245 (62.8) | 145 (37.2) |  |
|  | Three Years | 237 (64.0) | 133 (36.0) |  |
|  | Four Years | 272 (59.4) | 186 (40.6) |  |
| Education Household Head | No or Primary | 541 (62.3) | 328 (37.7) | < 0.001** |
|  | Secondary | 401 (72.9) | 149 (27.1) |  |
|  | Tertiary | 39 (92.1) | 3 (7.9) |  |
| Residence Type | Rural | 1052 (64.7) | 575 (35.3) | < 0.001* |
|  | Urban | 137 986.2) | 597 (13.8) |  |
| Province | Central | 71 (78.9) | 19 (21.1) | < 0.001** |
|  | Copperbelt | 150 (73.5) | 54 (26.5) |  |
|  | Eastern | 399 (76.0) | 126 (24.0) |  |
|  | Luapula | 121 (41.7) | 169 (58.3) |  |
|  | Lusaka | 57 (87.7) | 8 (12.3) |  |
|  | Muchinga | 75 (59.1) | 52 (40.9) |  |
|  | Northern | 88 (57.9) | 64 (42.1) |  |
|  | North-Western | 60 (62.5) | 36 (37.5) |  |
|  | Southern | 85 (97.7) | 2 (2.3) |  |
|  | Western | 83 (55.3) | 67 (44.7) |  |
| Wealth Quintiles | 1 (Lowest) | 215 (50.3) | 212 (49.7) | < 0.001* |
|  | 2 (Upper Low) | 199 (57.0) | 150 (43.0) |  |
|  | 3 (Middle) | 279 (73.0) | 103 (27.0) |  |
|  | 4 (Lower High) | 318 (75.5) | 103 (24.5) |  |
|  | 5 (Highest) | 178 (86.0) | 29 (14.0) |  |
| House Indoor Residual Spray | Sprayed | 1189 (66.6) | 597 (33.4) | < 0.001*** |
|  | Not Sprayed | 0 | 0 |  |
| Months Ago Sprayed | 0 - 3 Months | 185 (70.3) | 78 (29.7) | < 0.001* |
|  | 4 - 6 Months | 652 (61.6) | 407 (38.4) |  |
|  | 7 - 9 Months | 200 (69.2) | 89 (30.8) |  |
|  | 10 - 12 Months | 152 (86.9) | 23 (13.1) |  |
| ITN Use | Slept ITN | 558 (74.1) | 195 (25.90) | < 0.001* |
|  | Not Slept ITN | 92 (58.2) | 66 (41.8) |  |
| Haemoglobin | Anaemic | 491 (53.2) | 432 (46.8) | < 0.001* |
|  | Not Anaemic | 655 (81.3) | 151 (18.7) |  |
| Fever in last two weeks | Fever | 188 (44.8) | 232(55.2) | < 0.001* |
|  | No Fever | 1001 (73.3) | 365 (26.7) |  |

\* Chi-Square Test of Association
\*\* Fisher's Exact test
\*\*\* Test of Proportions

### Malaria Prevalence Ratios Adjusted for Confounders

The prevalence ratio of malaria among the four cohorts was significantly different in the univariate analysis, however, when adjusted for other confounders, the prevalence ratios were not statistically significant. This indicates that within one year, the protective effect of IRS under field conditions is not different. Other significant variables include age group, children who were either below one year or one year old were at less risk of malaria compared to those who are older. Those whose parents or guardian heads of households had secondary or tertiary education were also less at risk of malaria compared to those whose parents or guardian heads of households had no or just primary education. Other variables that were significantly associated with less risk of malaria include staying in urban areas compared to rural areas, staying in Eastern and Southern provinces compared to staying in Luapula province, being in the highest wealth quintile compared to being in the poorest quintile, not being anaemic and not having had fever during the survey. Table 3 summarises the unadjusted and adjusted prevalence ratios of malaria among the children in the four cohorts and other variables that were analysed in this study.

**Table 3:**
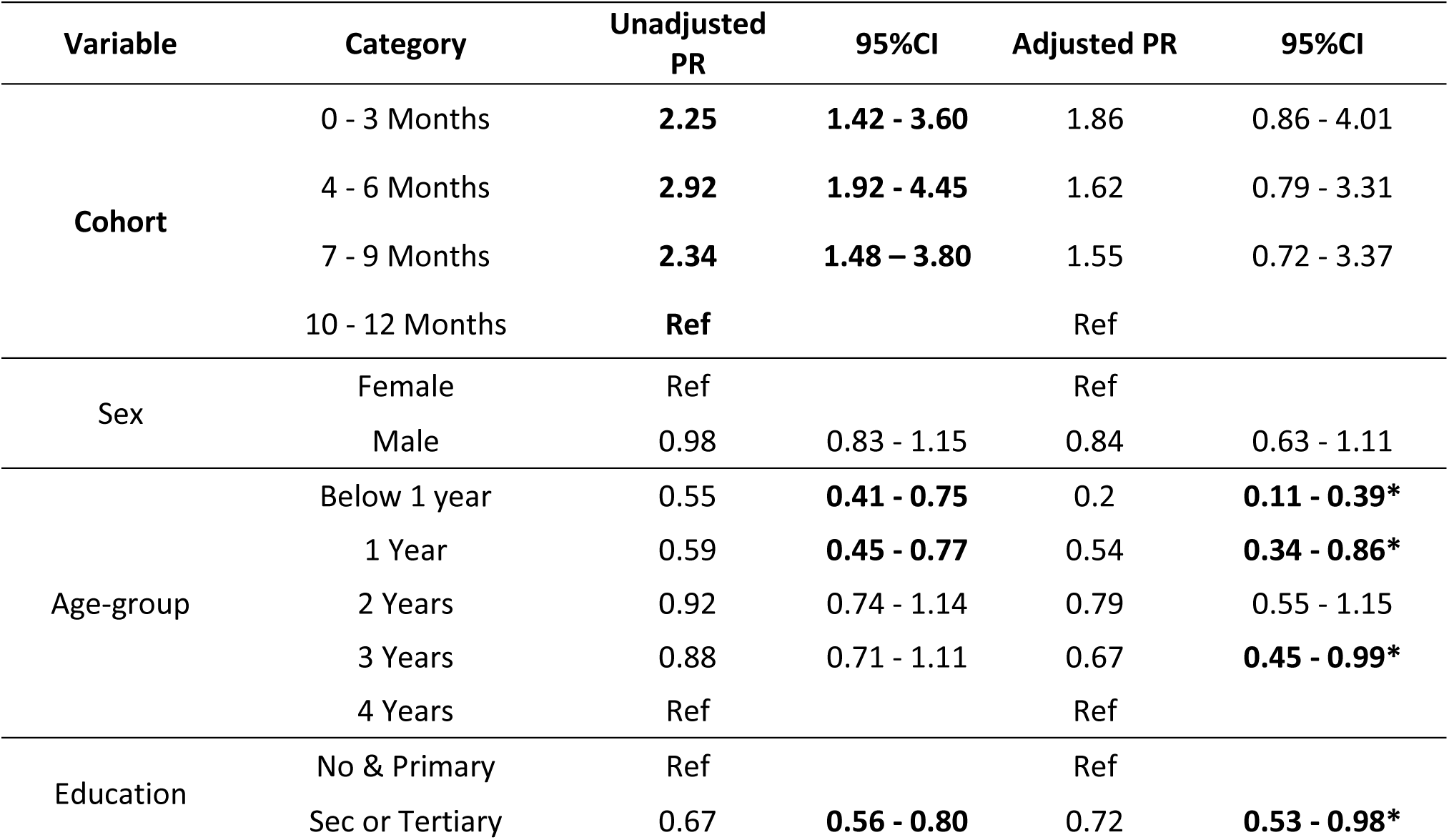
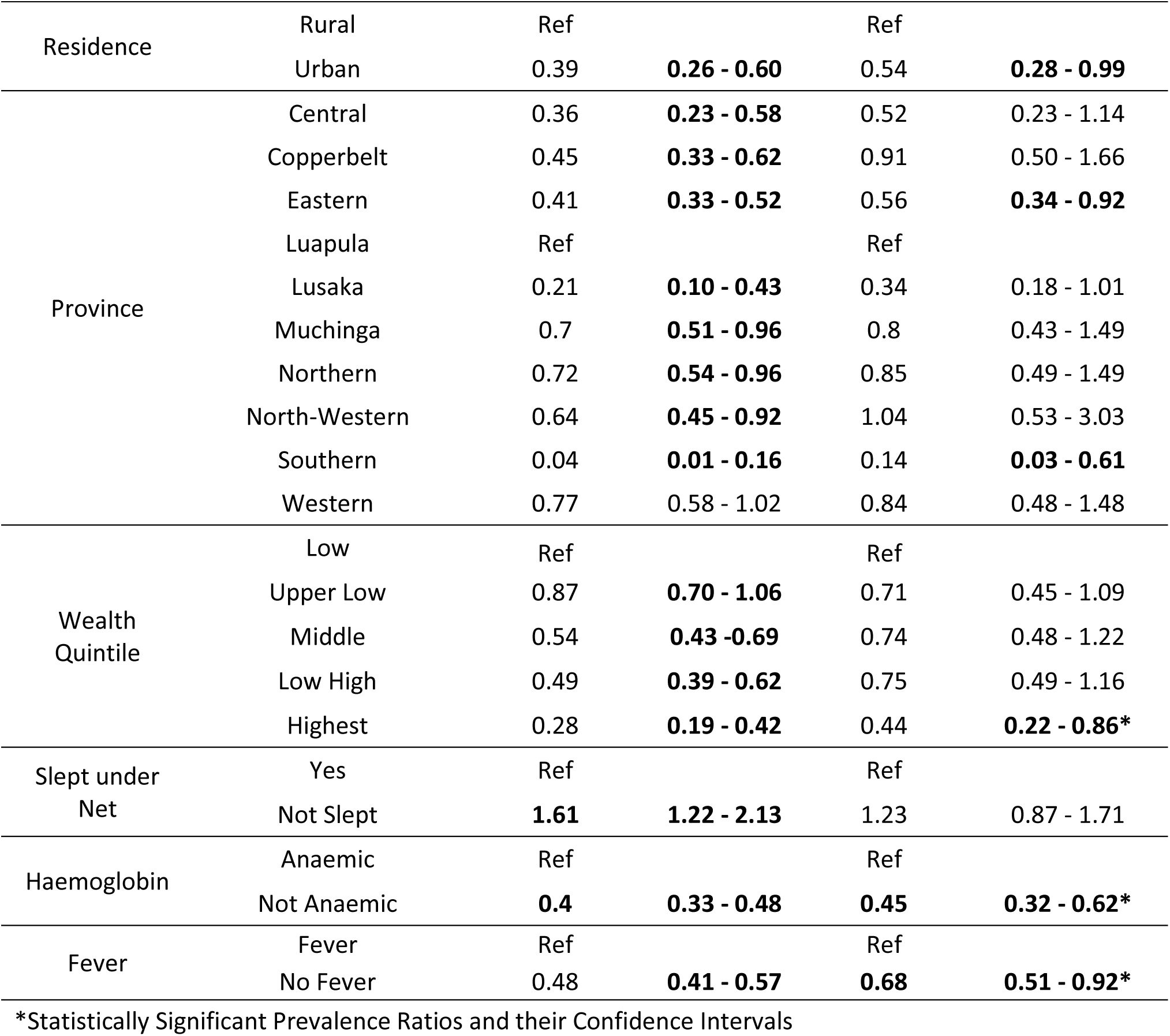
Unadjusted and Adjusted Prevalence Ratios for Malaria.

| Variable | Category | Unadjusted PR | 95%CI | Adjusted PR | 95%CI |
| --- | --- | --- | --- | --- | --- |
| **Cohort** | 0 - 3 Months | **2.25** | **1.42 - 3.60** | 1.86 | 0.86 - 4.01 |
|  | 4 - 6 Months | **2.92** | **1.92 - 4.45** | 1.62 | 0.79 - 3.31 |
|  | 7 - 9 Months | **2.34** | **1.48 – 3.80** | 1.55 | 0.72 - 3.37 |
|  | 10 - 12 Months | **Ref** |  | Ref |  |
| **Sex** | Female | Ref |  | Ref |  |
|  | Male | 0.98 | 0.83 - 1.15 | 0.84 | 0.63 - 1.11 |
| **Age-group** | Below 1 year | 0.55 | **0.41 - 0.75** | 0.2 | **0.11 - 0.39\*** |
|  | 1 Year | 0.59 | **0.45 - 0.77** | 0.54 | **0.34 - 0.86\*** |
|  | 2 Years | 0.92 | 0.74 - 1.14 | 0.79 | 0.55 - 1.15 |
|  | 3 Years | 0.88 | 0.71 - 1.11 | 0.67 | **0.45 - 0.99\*** |
|  | 4 Years | Ref |  | Ref |  |
| **Education** | No & Primary | Ref |  | Ref |  |
|  | Sec or Tertiary | 0.67 | **0.56 - 0.80** | 0.72 | **0.53 - 0.98\*** |
| Residence | Rural | Ref |  | Ref |  |
|  | Urban | 0.39 | **0.26 - 0.60** | 0.54 | **0.28 - 0.99** |
| Province | Central | 0.36 | **0.23 - 0.58** | 0.52 | 0.23 - 1.14 |
|  | Copperbelt | 0.45 | **0.33 - 0.62** | 0.91 | 0.50 - 1.66 |
|  | Eastern | 0.41 | **0.33 - 0.52** | 0.56 | **0.34 - 0.92** |
|  | Luapula | Ref |  | Ref |  |
|  | Lusaka | 0.21 | **0.10 - 0.43** | 0.34 | 0.18 - 1.01 |
|  | Muchinga | 0.7 | **0.51 - 0.96** | 0.8 | 0.43 - 1.49 |
|  | Northern | 0.72 | **0.54 - 0.96** | 0.85 | 0.49 - 1.49 |
|  | North-Western | 0.64 | **0.45 - 0.92** | 1.04 | 0.53 - 3.03 |
|  | Southern | 0.04 | **0.01 - 0.16** | 0.14 | **0.03 - 0.61** |
|  | Western | 0.77 | 0.58 - 1.02 | 0.84 | 0.48 - 1.48 |
| Wealth Quintile | Low | Ref |  | Ref |  |
|  | Upper Low | 0.87 | **0.70 - 1.06** | 0.71 | 0.45 - 1.09 |
|  | Middle | 0.54 | **0.43 - 0.69** | 0.74 | 0.48 - 1.22 |
|  | Low High | 0.49 | **0.39 - 0.62** | 0.75 | 0.49 - 1.16 |
|  | Highest | 0.28 | **0.19 - 0.42** | 0.44 | **0.22 - 0.86\*** |
| Slept under Net | Yes | Ref |  | Ref |  |
|  | Not Slept | **1.61** | **1.22 - 2.13** | 1.23 | 0.87 - 1.71 |
| Haemoglobin | Anaemic | Ref |  | Ref |  |
|  | Not Anaemic | **0.4** | **0.33 - 0.48** | **0.45** | **0.32 - 0.62\*** |
| Fever | Fever | Ref |  | Ref |  |
|  | No Fever | 0.48 | **0.41 - 0.57** | **0.68** | **0.51 - 0.92\*** |

\*Statistically Significant Prevalence Ratios and their Confidence Intervals

## DISCUSSION

This study set out to find the prevalence ratios of malaria infections among children whose households were sprayed at different times during the preceding 12 months of the Malaria Indicator Survey of 2021 in Zambia. The study was, therefore, able to construct four three monthly cohorts over the 12 months and compared the malaria prevalence ratios. Households that are sprayed are usually stuck with a sticker that specifies the date and chemical used by IRS. The study found no significant differences in the malaria prevalence ratios for the zero to three months, four to six months, seven to nine months and ten to twelve months cohorts. These findings imply that the protective effects of IRS in Zambia within the first twelve months under field conditions were comparable and there were no significant statistical differences.

There is limited literature on studies comparing malaria prevalence using months after IRS, many studies on IRS compare different chemicals or IRS intervention versus no intervention (10, 32, 33). Other studies have also looked at integrating IRS with other interventions like ITNs, house modification, larvicides and repellent sprays (33). This study is therefore unique as it adds to the body of knowledge particularly from field settings that look at comparing malaria prevalence months after spraying with residual chemicals. Based on the Zambian guidelines in the National Malaria Elimination Plan 2017 to 2021 which was in operation then, the country uses mainly Pirimiphos methyl and Fludora fusion which are known to be effective and the vectors are currently not resistant to unlike carbamate and pyrethroids (18, 34, 35). Further, the two groups of chemicals are also known to have long-lasting effectiveness of about eight to twelve months (18). Finding no differences in the different month periods does not imply that IRS is not effective, but rather that the differences in malaria prevalence among sprayed households for different month cohorts were not statistically significant. Recent studies in Zambia found that IRS was effective in reducing malaria incidence and prevalence among children in sprayed households (9, 36). Studies done elsewhere in Africa have also found that IRS is effective in preventing malaria (10).

While the study focus was on IRS, it also found other factors to be significantly associated with malaria prevalence reduction, younger children below one year were associated with less risk of malaria compared to older children. This has been corroborated by other studies which found that infants were at less risk of malaria compared to older children (37). Infants tend to still retain maternal antibodies against malaria during birth and also during breastfeeding, further, the level of care during infancy is more compared to older children (37). Other significant factors include higher education compared to no or primary education among parents and guardians, wealthier households compared to poor households and residing in urban areas compared to rural areas. These were also affirmed by earlier studies, so they are not unique to this study (38). Being anaemic and having a fever were associated with having a higher risk of malaria compared to not being anaemic and not having a fever respectively. Equally, other studies have reported similar findings (39).

## CONCLUSION

This study found that over a period of one year, the protective effect of Indoor Residual Spraying was comparable among under-five children for the zero to three months, four to six months, seven to nine months and ten to twelve months cohorts. Factors that were associated with malaria prevalence reduction included children below one year, more educated and wealthy guardians or parents, residing in urban areas, eastern and southern provinces of Zambia, and not having anaemia or fever.

### Recommendations

Based on the findings of this study, the authors recommend IRS not only be at the beginning of the rainy season in Zambia as the current recommendations and practices have been. Usually, the logistics of procuring and distributing IRS chemicals are delayed by procurement processes and transporting such that spraying is done during the actual rainy season. Some residents refuse to have their houses to be sprayed because of the fear that their household properties can be spoiled by the rains during the rainy season. The author therefore recommends more research especially prospective studies such as prospective cohorts and randomised controlled trials which if affirmed can change policies to allow IRS throughout the year with long-acting chemicals such as Pirimiphos methyl and Fludora fusion to increase the acceptability of IRS among residents during off season.

### Limitations

The study used secondary data from a nationwide cross-sectional survey and the measurement of malaria was done only once during the rainy season. Malaria dynamics change during the rainy season and dry season, therefore the findings of this study may not be generalizable to the dry season. Further, the design of the primary study which was cross-sectional cannot confer causal effects, our findings can therefore only be confined to associations.

## Data Availability statement

The data is available upon request through the ministry of Health, Zambia

